# Cellular and humoral immune response to a third dose of BNT162b2 COVID-19 vaccine – a prospective observational study

**DOI:** 10.1101/2022.03.10.22272204

**Authors:** Jonas Herzberg, Bastian Fischer, Heiko Becher, Ann-Kristin Becker, Human Honarpisheh, Salman Yousuf Guraya, Tim Strate, Cornelius Knabbe

**Affiliations:** Department of Surgery – Krankenhaus Reinbek St. Adolf-Stift, Hamburger Strasse 41, 21465 Reinbek, Germany; Institut für Laboratoriums- und Transfusionsmedizin, Herz- und Diabeteszentrum NRW, Georgstraße 11, 32545 Bad Oeynhausen, Germany; Institute of Medical Biometry and Epidemiology, University Medical Center Hamburg-Eppendorf, Martinistrasse 52, 20246 Hamburg, Germany; Asklepios Klinik Harburg, Abteilung für Psychiatrie und Psychotherapie, Eißendorfer Pferdeweg 52, 21075 Hamburg, Germany; Clinical Sciences Department, College of Medicine, University of Sharjah, P. O. Box 27272 Sharjah, United Arab Emirates

**Keywords:** SARS-CoV-2, humoral and cellular immunity, Health Care worker, Seroprevalence, Vaccination, BNT162b2, COVID-19, third dose, boosting, immunity

## Abstract

**Background:** Since the introduction of various vaccines against SARS-CoV-2 at the end of 2020, rates of infection have continued to climb worldwide. This led to the establishment of a third dose vaccination in several countries, known as a booster. To date, there has been little real-world data about the immunological effect of this strategy.

**Methods:** We compared the humoral- and cellular immune response before and after the third dose of BioNTech/Pfizer vaccine BNT162b2, following different prime-boost regimes. Humoral immunity was assessed by determining anti-SARS-CoV-2 binding antibodies using a standardized quantitative assay. In addition, neutralizing antibodies were measured using a commercial surrogate ELISA-assay. Interferon-gamma release was measured after stimulating blood-cells with SARS-CoV-2 specific peptides using a commercial assay to evaluate the cellular immune response.

**Results:** The median antibody level increased significantly after the third dose to 2663.1 BAU/ml vs. 101.4 BAU/ml (p < 0.001) before administration of the boosting dose. This was also detected for neutralizing antibodies with a binding inhibition of 99.68% ± 0.36% vs. 69.06% ± 19.88% after the second dose (p < 0.001).

96.3% of the participants showed a detectable T-cell-response after the third dose with a mean interferon-gamma level of 2207.07 mIU/ml ± 1905 mIU/ml.

**Conclusion:** This study detected a BMI-dependent increase after the third dose of BNT162b2 following different vaccination protocols, whereas all participants showed a significant increase of their immune response. This, in combination with the limited post-vaccination-symptoms underlines the potential beneficial effect of a BNT162b2-boosting dose.

## Introduction

Following an initially flattened curve of COVID-19 infections, due to vaccination campaigns all over the world, there came a subsequent resurgence of COVID-19 infections worldwide [1]. New variants of SARS-CoV-2 and decreasing immunity after vaccination over time [2,3], have caused an increased rate of infection and hospitalization also in vaccinated people [1]. A well-established approach to handle the so-called secondary vaccine failure, is the use of a booster dose – an additional vaccine dose after the initial round of immunization [1,4]. The effectiveness of a third dose of BioNTech/Pfizer vaccine was suggested by different studies [5–7]. Therefore, after approval by the regulatory authorities in US [8] and in the European Union [9], several countries such as Israel, USA, UK and Germany, initiated a vaccination program for administration of a third dose in vulnerable groups at least 5 months after complete vaccination [1,10].

The effect of this third dose on the immune response and the resulting protective effects have been analyzed in register or clinical studies [1,7,11,12].

As the impact of different vaccines on the immune system varies, considering both cellular and humoral immunity is crucial. The detected levels of neutralizing antibodies have been shown to be higher after administration of the BioNTech/Pfizer vaccine BNT162b2 than those after receiving the AstraZeneca vaccine ChAdOx1, although this could not be shown for T-cell responses [13]. These differences have led to the discussion regarding potential benefit from a heterologous vaccination strategy [13–16].

We aimed to evaluate the humoral- and cellular immune response after a third booster-dose of BNT162b2 following an initial administration of either two doses of BNT162b2 (Group 1), two doses of ChAdOx1 (Group 2) or cross-vaccination of BNT162b2+ChAdOx1 (Group 3) in a real-world setting analyzing one of the most important groups in this global pandemic – health care workers.

## Methods

### Study design

In this study we evaluated the effect of administration of a third dose of BioNTech/Pfizer mRNA vaccine BNT162b2, within a longitudinal study in health care workers initiated in April 2020 [17]. All employees of the Hospital Reinbek St. Adolf-Stift, a secondary care hospital located in Northern Germany, were eligible to participate in the study. The vaccination program for employees was established in December 2020. At the end of October 2021, the Robert-Koch-Institute recommended administration of a third dose in specific groups [18]. Following this recommendation, beginning in November 2021 all employees who had received their second vaccination more than 6 months prior, were invited for a third booster-dose. In order to have a reference value for the determination of the booster-effect, blood samples were initially collected and analyzed before the administration of the third vaccine dose on November 13^th^ – 14^th^ 2021 [2].

To evaluate the efficiency of the third dose all participants were asked to provide an additional blood sample and complete a questionnaire 4 weeks after the booster-vaccination (December 13^th^ – 14^th^ 2021).

### Anti-SARS-CoV-2-IgG antibodies

The anti-SARS-CoV-2-igG antibody-titer was expressed in Binding Antibody Units per ml (BAU/ml) to stay in accordance with the WHO standard. Antibodies were determined by using a fully automated quantitative anti-SARS-CoV-2-assay (IgG) from Abbott (Chicago, USA). As recommended by the manufacturer, a value below 7.1 BAU/ml was determined to be negative whereas values ≥7.1 BAU/ml were determined to be positive.

### Neutralizing antibodies against SARS-CoV-2

To evaluate the neutralizing anti-SARS-CoV-2 antibodies, all samples were additionally tested using the NeutraLISA™ SARS-CoV-2 Neutralization Antibody Detection KIT (Euroimmun, Lübeck, Germany). Results were presented in binding inhibition, where values ≥ 35 % were considered positive.

### T-cell-response

In addition to the humoral immune response, the cellular immunity to SARS-CoV-2 was also assessed, using the Quan-T-cell SARS-CoV-2 kit (Euroimmung, Lübeck, Germany) working as an Interferon-gamma release assay (IGRA). The analysis for this Quan-T-cell test was performed as previously reported [2]. IFN-gamma concentration was expressed as mIU/ml. Values ≥200 mIU/ml were considered positive.

### Questionnaire

Before and after administration of the third dose of vaccine, all participants were asked to complete a questionnaire regarding their prior vaccinations, smoking behaviors and especially after the third dose, possible side effects of the third dose. Specifically, participants were questioned about localized symptoms (local pain, lymphadenopathy) and systemic symptoms, such as fever or fatigue.

This study was conducted in accordance with the Declaration of Helsinki. All participants provided written and informed consent before inclusion. This study was prospectively registered at the German Clinical Trial Register (DRKS00021270) after approval by the Ethics Committee of the Medical Association Schleswig-Holstein.

### Statistical analysis

All variables are presented as means or medians with standard deviation or interquartile range. Categorical variables are shown as numbers with percentages. Statistical analysis was made using IBM SPSS Statistics Version 25 (IBM Co., Armonk, NY, USA). In addition, GraphPad Prism 9 and IBM SPSS Statistics Version 25 (IBM Co., Armonk, NY, USA) were used for graphics.

Relationships between categorial variables were tested using the Chi-square test or the Fisher’s exact test, depending on the size of the groups. Inter-group differences were analyzed using Mann-Whitney-U test or Kruskal-Wallis-test reporting the mean with standard deviation. Pre- and post-third dose antibody and INF-gamma levels were compared using Wilcoxon sign rank test. A p-value < 0.05 was considered statistically significant. To investigate the joint effect of age, sex, body mass index and current smoking on the relative increase of antibody-level, a linear regression analysis was done to take the antibody level before booster into account. The IgG-levels had a skewed distribution and was logarithmized for the regression analysis yielding an approximately normal distribution. As the dependent variable, the difference of the logarithmized IgG-levels after and before booster was used. The backward selection method was used to determine the final model.

## Results

Before administration of the third vaccine-dose, 310 participants provided a blood specimen, of which 243 resubmitted a blood sample and completed questionnaire exactly 4 weeks after the booster dose (follow-up-rate 78.39%).

The participants included 59 (24.3%) male and 184 (75.7%) female participants with a mean age of 46.33 ± 11.44 years. The characteristics of the study cohort are described in **Table 1**, grouped according to their initial vaccination protocol.

**Table 1:** Characteristics of the study cohort (n = 243).

|  | **All**<br>**(n = 243)** | **Group 1**<br>**(n = 179)** | **Group2**<br>**(n = 50)** | **Group 3**<br>**(n = 12)** |
| --- | --- | --- | --- | --- |
| **Age, M ± SD [years]** | 46.33 ±<br>11.44 | 46.52 ±<br>10.99 | 43.94 ±<br>12.13 | 51.50 ±<br>13.62 |
| **Sex, n (%)** |  |  |  |  |
| **Male** | 59 (24.3) | 45 (25.1) | 11 (22.0) | 2 (16.7) |
| **Female** | 184 (75.7) | 134 (74.9) | 39 (78.0) | 10 (83.3) |
| **BMI, M ± SD [kg/m<sup>2</sup>]** | 25.95 ±<br>5.02 | 26.00 ±<br>5.08 | 26.47 ± 5.09 | 23.57 ±<br>3.20 |
| **Smoking, n (%)** | 56 (23.0) | 47 (26.3) | 8 (16.0) | 1 (8.3) |
| **Comorbidities, n (%)** |  |  |  |  |
| **Cardiac** | 44 (18.1) | 33 (18.4) | 8 (16.0) | 3 (25.0) |
| **Pulmonary** | 20 (8.2) | 13 (7.3) | 5 (10.0) | 2 (16.7) |
| **Metabolic** | 39 (16.0) | 30 (16.8) | 7 (14.0) | 2 (16.7) |
| **Immunologic** | 7 (2.9) | 6 (3.4) | 1 (2.0) | 0 (0) |
| **Other** | 50 (20.6) | 39 (21.8) | 8 (16.0) | 3 (25.0) |
| **Reaction after booster,**<br>**n (%)** | 189 (77.8) | 139 (77.7) | 40 (80.0) | 8 (66.7) |
| **Local pain** | 147 (60.5) | 109 (60.9) | 31 (62.0) | 5 (41.7) |
| **Headache** | 49 (20.2) | 32 (17.9) | 13 (26.0) | 4 (33.3) |
| **Fatigue** | 91 (37.4) | 61 (34.1) | 26 (52.0) | 3 (25.0) |
| **Fever** | 30 (12.3) | 21 (11.7) | 9 (18.0) | 0 (0) |
| **Limb pain** | 47 (19.3) | 34 (19.0) | 10 (20.0) | 3 (25.0) |
| **Lymphadenopathy** | 12 (4.9) | 11 (6.1) | 0 (0) | 1 (8.3) |

All participants received a booster dose of BNT162b2, irrespective of their primary vaccination protocol. The participants included 179 (73.7%) individuals who had previously received two doses of BNT162b2 (Group 1), 50 (20.6%) individuals who had received heterologous vaccinations with BNT162b2+ChAdOx1 (Group 2), 12 (4.9%) individuals who were double vaccinated with ChAdOx1 (Group 3), and 2 (0.8%) individuals who had a natural SARS-CoV-2 infection followed by a single dose of BNT162b2.

### Whole study cohort

#### Anti SARS-CoV-2-IgG binding antibodies

Prior to the booster dose, all participants still showed an antibody level above the manufacturer’s cutoff (>7.1 BAU/ml). The median antibody level increased significantly after the third dose, when the whole study cohort was considered (2663.1, IQR 1700.7-4180.9 vs. 101.4, IQR 60.6-163.6; p < 0.001). All individuals showed increased antibody-levels with a median increase of 2539.55, IQR 1613.2-4002.5 (**Figure 1A**). The median relative increase was 25.9 (IQR 15.5-48.7).

**Figure 1:**
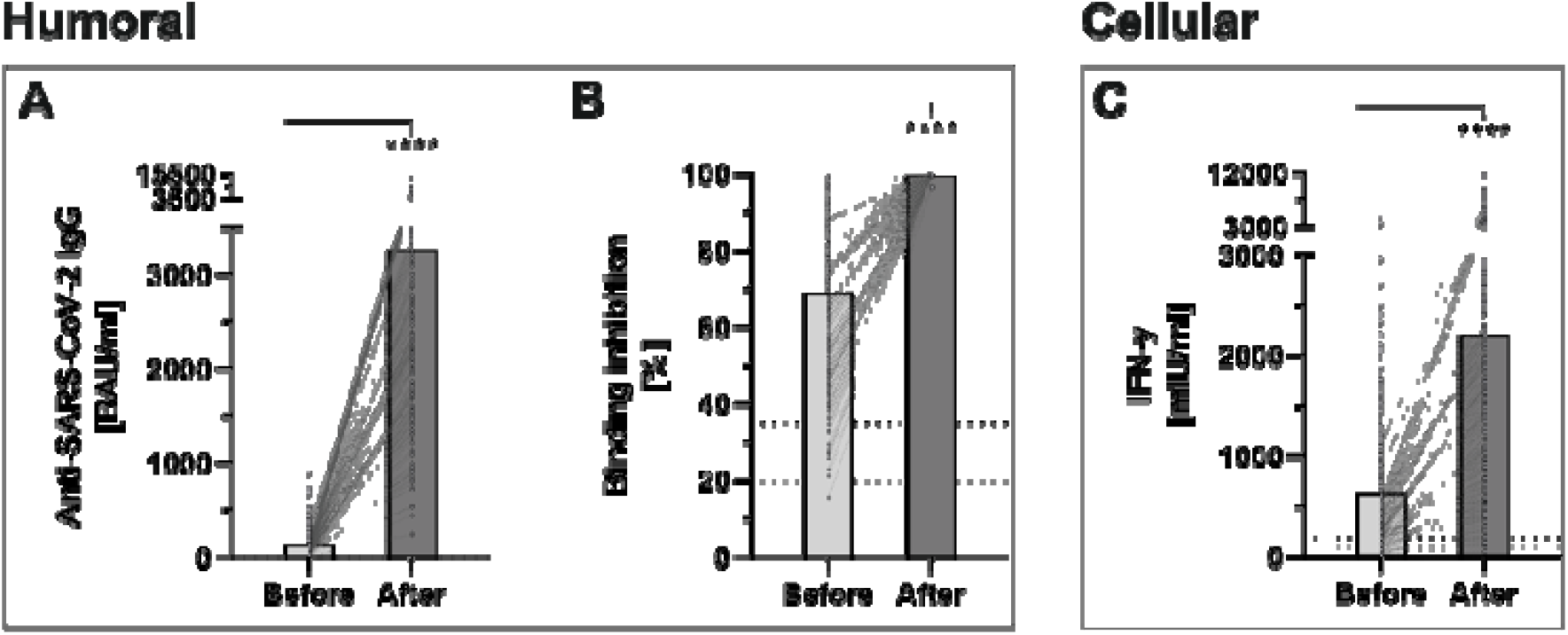
Determination of humoral- and cellular immunity against-SARS-CoV-2 before and four weeks after third booster vaccination with BNT162b2 (n = 243). **A**: Anti-SARS-CoV-2-IgG binding antibodies were determined using a quantitative assay from Abbott. In keeping with the WHO-standard, data were expressed in Binding Antibody Units per ml (BAU/ml). Samples were marked seronegative below 7.1 BAU/ml whereas values above 7.1 BAU/ml were determined to be positive, as mentioned by the manufacturer. **B**: Binding inhibition capability of Neutralizing anti-SARS-CoV-2 antibodies was determined using the NeutraLISA™ SARS-CoV-2 Neutralization Antibody Detection KIT from Euroimmun. According to the manufacturer, binding inhibition values above 35% were considered positive (horizontal black dotted line), whereas values between 20% and 35% were considered equivocal (horizontal gray dotted line). **C**: Cellular immunity to SARS-CoV-2 was assessed by using an Interferon (IFN)-gamma release assay (IGRA) from Euroimmun (Quan-T-cell SARS-CoV-2 kit). Values > 200 mIU/ml were considered positive (horizontal black dotted line), whereas values between 100-200 mIU/ml were considered equivocal (horizontal gray dotted line). ****p < 0.0001 (Mann-Whitney U-test).

#### Neutralizing antibodies

The administration of a third dose of BNT162b2 vaccine also caused a significant increase in neutralizing antibodies, throughout all study groups (99.68 % ± 0.36 % vs. 69.06 % ± 19.88 %; p < 0.001) (**Figure 1B**). No participants showed a binding inhibition capability below 96 % following the booster-vaccination.

#### T-cell response

Before the booster dose in November 2021, 73.4 % of the participants still had a detectable T-cell response. This rate increased in the evaluation after the third dose to 96.3 %, with a significantly higher mean INF-gamma level (2207.07 ± 1905.55 vs. 630.21 ± 650.53; p < 0.001) (**Figure 1C**).

### Vaccination-strategy specific cohorts

#### Anti SARS-CoV-2 binding antibodies

Anti-SARS-CoV-2 binding antibody levels differed significantly between the subgroups (representing different prior vaccination protocols), prior to booster-administration. In brief, pre-boost antibody levels were highest in Group 2 (183.1 BAU/ml), followed by Group 1 (124.9 BAU/ml) and Group 3 (52.75 BAU/ml). The administration of a third BNT162b2 dose led to a highly significantly increased antibody-level, across all analyzed subgroups. Inductive effects were highest within Group 3 participants (mean 2358 vs. 52.75 (4,370%), p = 0.002), followed by Group 1 (mean 3661 vs. 124.9 (2,831 %), p < 0.001) and Group 2 (mean 2122 vs. 183.1 (1,058%), p < 0.001) participants (**Figure 2** and **Table 2**).

**Table 2:** Values in accordance to the immunization protocol.

|  | All |  |  | Group 1 |  |  | Group 2 |  |  | Group 3 |  |  |
| --- | --- | --- | --- | --- | --- | --- | --- | --- | --- | --- | --- | --- |
|  | IgG<br>[BAU/ml] | INF-y<br>[mIU/ml] | NAK<br>[%] | IgG<br>[BAU/ml] | INF-y<br>[mIU/ml] | NAK<br>[%] | IgG<br>[BAU/ml] | INF-y<br>[mIU/ml] | NAK<br>[%] | IgG<br>[BAU/ml] | INF-y<br>[mIU/ml] | NAK<br>[%] |
| **All** | 3266.26 ±<br>2194.17 | 2207.07 ±<br>1905.55 | 99.68 ±<br>0.36 | 3660.88 ±<br>2250.13 | 2013.70 ±<br>1780.13 | 99.71 ±<br>0.28 | 2357.80 ±<br>1815.63 | 3090.58 ±<br>2671.98 | 99.70 ±<br>0.18 | 2122.32 ±<br>1572.44 | 2424.93 ±<br>1520.45 | 99.59 ±<br>0.57 |
| **Sex** |  |  |  |  |  |  |  |  |  |  |  |  |
| **Male** | 3397.52 ±<br>2675.29 | 2010.27 ±<br>1526.05 | 99.65 ±<br>0.52 | 4064.25 ±<br>2716.71 | 2130.64 ±<br>1527.94 | 99.74 ±<br>0.16 | 1032.35 ±<br>707.32 | 771.40 ±<br>274.99 | 99.51 ±<br>0.28 | 1232.95 ±<br>707.32 | 1428.37 ±<br>1133.33 | 99.31 ±<br>1.13 |
| **Female** | 3224.17 ±<br>2022.59 | 2270.53 ±<br>2012.33 | 99.69 ±<br>0.29 | 3525.42 ±<br>2064.21 | 1974.13 ±<br>1861.38 | 99.70 ±<br>0.31 | 2622.89 ±<br>1872.16 | 3554.42 ±<br>2698.76 | 99.73 ±<br>0.14 | 2373.17 ±<br>1662.26 | 2706.01 ±<br>1508.11 | 99.66 ±<br>0.23 |
| **Smoking** |  |  |  |  |  |  |  |  |  |  |  |  |
| **Yes** | 3419.37 ±<br>2328.14 | 1649.21 ±<br>1347.08 | 99.71 ±<br>0.22 | 3737.61 ±<br>2390.63 | 1545.40 ±<br>1338.35 | 99.74 ±<br>0.12 | 2156.10 | 3934.84 | 99.61 | 1707.60 ±<br>880.06 | 1973.41 ±<br>1244.33 | 99.50 ±<br>0.47 |
| **No** | 3220.41 ±<br>2156.84 | 2375.03 ±<br>2016.98 | 99.68 ±<br>0.39 | 3633.56 ±<br>2206.79 | 2181.71 ±<br>1890.24 | 99.70 ±<br>0.32 | 2376.14 ±<br>1903.08 | 3013.83 ±<br>2788.49 | 99.70 ±<br>0.18 | 2201.32 ±<br>1668.18 | 2510.93 ±<br>1565.61 | 99.60 ±<br>0.59 |
| **Obesity** |  |  |  |  |  |  |  |  |  |  |  |  |
| **Yes** | 3772.66 ±<br>2586.61 | 1805.01 ±<br>1499.21 | 99.72 ±<br>0.10 | 4159.60 ±<br>2769.73 | 1501.28 ±<br>1307.93 | 99.72 ±<br>0.10 | N/A | N/A | N/A | 2760.68 ±<br>1737.00 | 2599.36 ±<br>1722.37 | 99.73 ±<br>0.09 |
| **No** | 3144.83 ±<br>2078.43 | 2303.98 ±<br>1982.24 | 99.67 ±<br>0.39 | 3543.94 ±<br>2104.28 | 2134.69 ±<br>1857.52 | 99.71 ±<br>0.31 | 2357.80 ±<br>1815.63 | 3090.58 ±<br>2671.98 | 99.70 ±<br>0.18 | 1898.04 ±<br>1469.99 | 2363.64 ±<br>1463.86 | 99.53 ±<br>0.66 |
| **Symptoms after vaccination** |  |  |  |  |  |  |  |  |  |  |  |  |
| **Yes** | 3397.22 ±<br>2277.30 | 2394.65 ±<br>1970.57 | 99.67 ±<br>0.39 | 3787.36 ±<br>2347.85 | 2203.38 ±<br>1891.74 | 99.70 ±<br>0.31 | 2768.40 ±<br>1962.11 | 3008.23 ±<br>1952.78 | 99.70 ±<br>0.18 | 2237.24 ±<br>1642.77 | 2622.83 ±<br>1601.94 | 99.57 ±<br>0.62 |
| **No** | 2807.92 ±<br>1819.79 | 1538.16 ±<br>1485.19 | 99.71 ±<br>0.18 | 3221.36 ±<br>1829.71 | 1337.65 ±<br>1078.61 | 99.73 ±<br>0.12 | 1536.60 ±<br>1324.94 | 3255.29 ±<br>4150.42 | 99.70 ±<br>0.18 | 1662.66 ±<br>1214.08 | 1633.30 ±<br>771.86 | 99.66 ±<br>0.33 |

**Figure 2:**
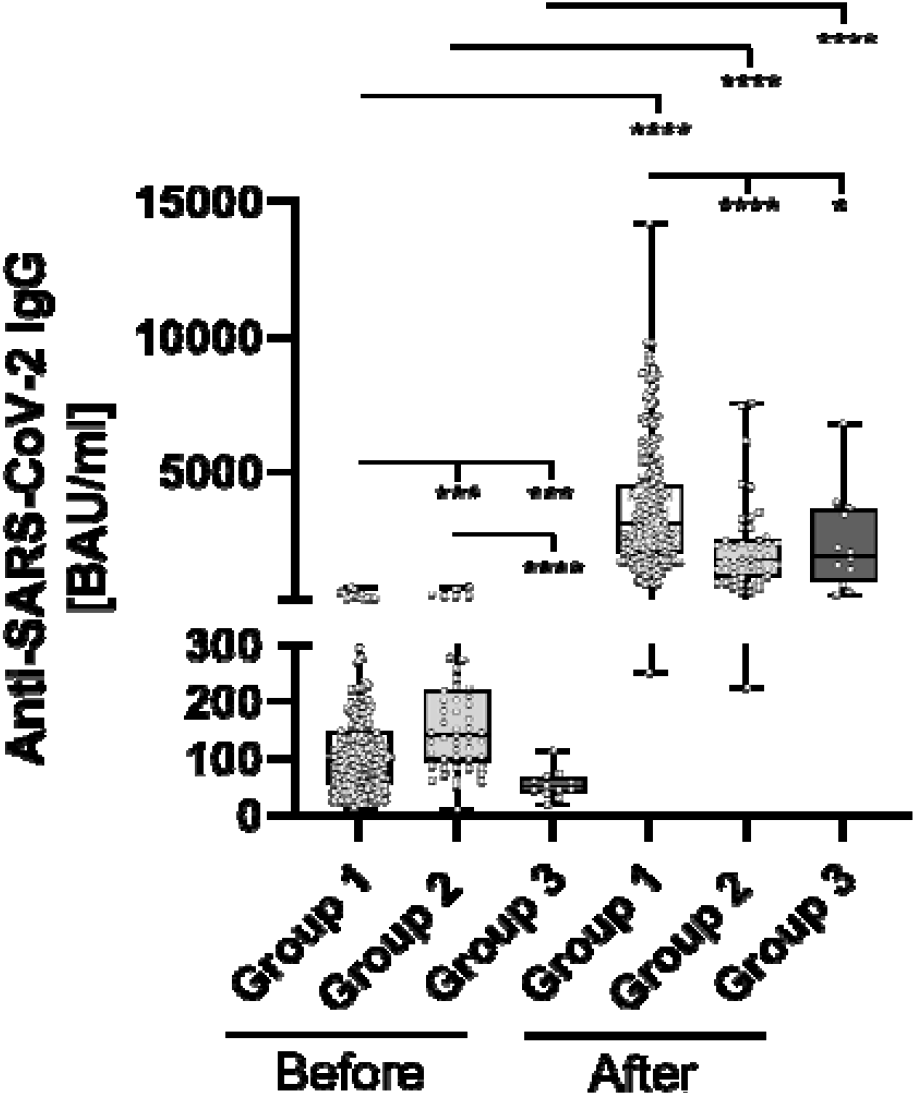
Comparative determination of anti SARS-CoV-2 IgG binding antibodies before and after third booster vaccination with BNT162b2. To evaluate differences between vaccination-strategies, participants were grouped into 3 cohorts: Group 1: three vaccine-doses of BNT162b2; Group 2: initially two vaccine-doses of ChAdOx1 and BNT162b2 booster-dosage; Group 3: heterologous vaccination-protocol (ChAdOx1+ BNT162b2) and BNT162b2 booster-dosage. Anti-SARS-CoV-2-IgG binding antibodies were determined before- and 4 weeks after third booster-dosage using a quantitative assay from Abbott. In keeping with the WHO-standard, data were expressed in Binding Antibody Units per ml (BAU/ml). Samples were marked seronegative below 7.1 BAU/ml whereas values above 7.1 BAU/ml were determined to be positive, as mentioned by the manufacturer. *p < 0.05; ***p < 0.001; ****p < 0.0001 (Mann-Whitney U-test).

#### Neutralizing antibodies

Neutralizing antibody levels also differed significantly between the groups prior to booster-administration (**Figure 3** and **Table 2**). Specifically, individuals within Group 3 showed the lowest percentage binding inhibition (49.55%), followed by Group 1 (67.66%) and Group 2 (77.57%) participants. The administration of a third BNT162b2 dose caused a significant induction of binding antibody capability within all three groups, whereby the strongest effect was detected for Group 3 (99.70% vs. 49.55% (induction: 101.21%)), followed by Group 1 (99.71% vs. 67.66% (induction: 47.37%)) and Group 2 (99.59% vs. 77.57% (induction: 28.39%)) participants. Post-booster binding inhibition capabilities did not differ significantly between the different groups.

**Figure 3:**
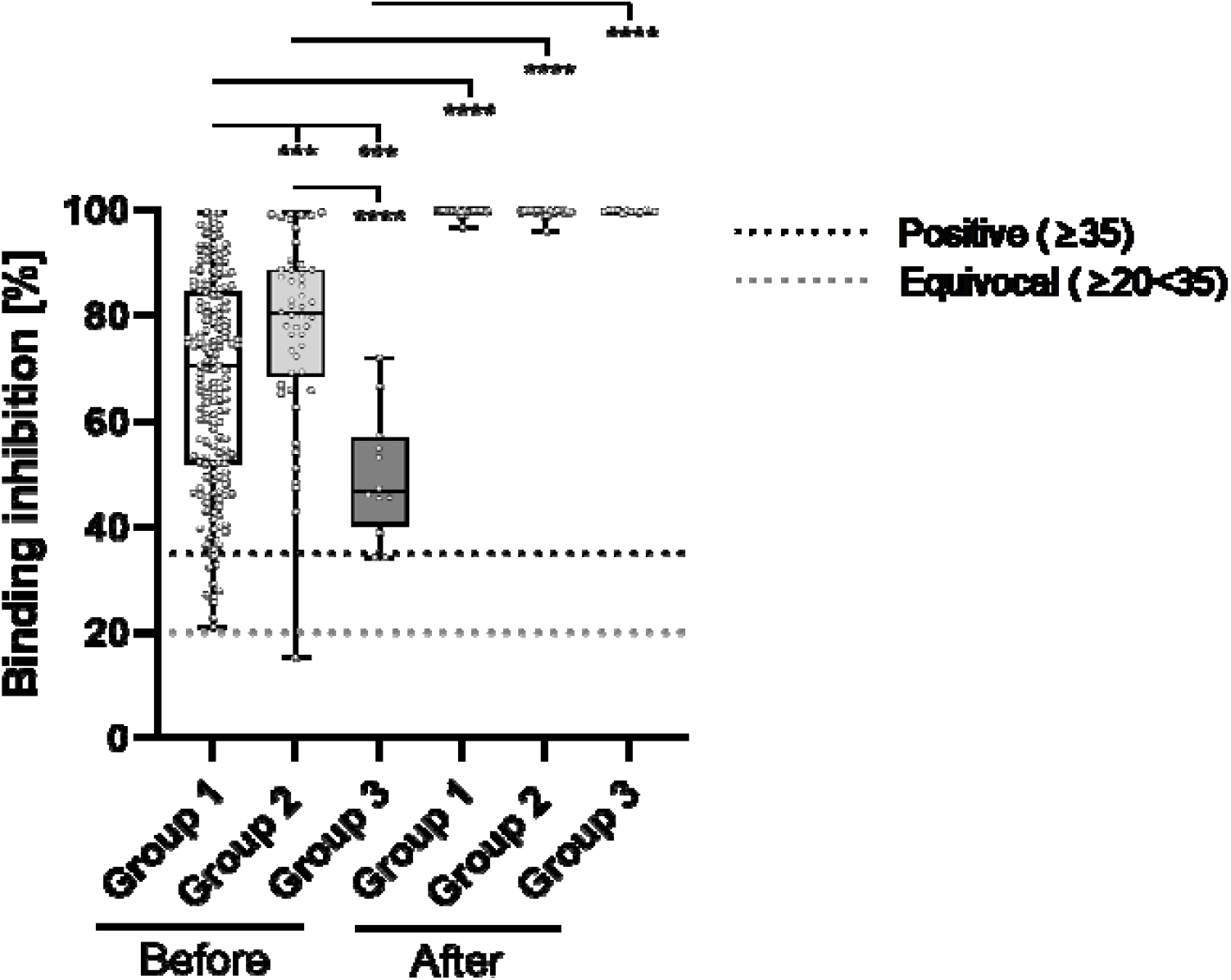
Comparative determination of neutralizing antibody binding-inhibition-capability before and after third booster vaccination with BNT162b2. To evaluate differences between vaccination-strategies, participants were grouped into 3 cohorts: Group 1: three vaccine-doses of BNT162b2; Group 2: initially two vaccine-doses of ChAdOx1 and BNT162b2 booster-dosage; Group 3: heterologous vaccination-protocol (ChAdOx1+ BNT162b2) and BNT162b2 booster-dosage. Binding inhibition capability of neutralizing anti-SARS-CoV-2 antibodies was determined using the NeutraLISA™ SARS-CoV-2 Neutralization Antibody Detection KIT from Euroimmun. According to the manufacturer, binding inhibition values above 35% were considered positive (horizontal black dotted line), whereas values between 20% and 35% were considered equivocal (horizontal gray dotted line). ***p< 0.001; ****p < 0.0001 (Mann-Whitney U-test).

#### T-cell response

Prior to the administration of a booster BNT162b2 dosage, SARS-CoV-2 specific IFN-gamma release of stimulated blood-cells significantly differed between individuals of each considered group (**Figure 4, Table 2**). Briefly, Group 1 participants showed the lowest IFN-gamma mean-value (560.4 mIU/ml), followed by individuals belonging to Group 3 (654.7 mIU/ml) and Group 2 (828.7 mIU/ml). SARS-CoV-2 specific t-cell response significantly increased in boostered individuals within all groups. The strongest percentage inductive effects were observed for Group 3 individuals (3091 vs. 654.7 (372.12%)), followed by participants belonging to group 1 (2014 vs. 560.4 (259.39%)) and group 2 (2425 vs. 828.7 (192.63%)).

**Figure 4:**
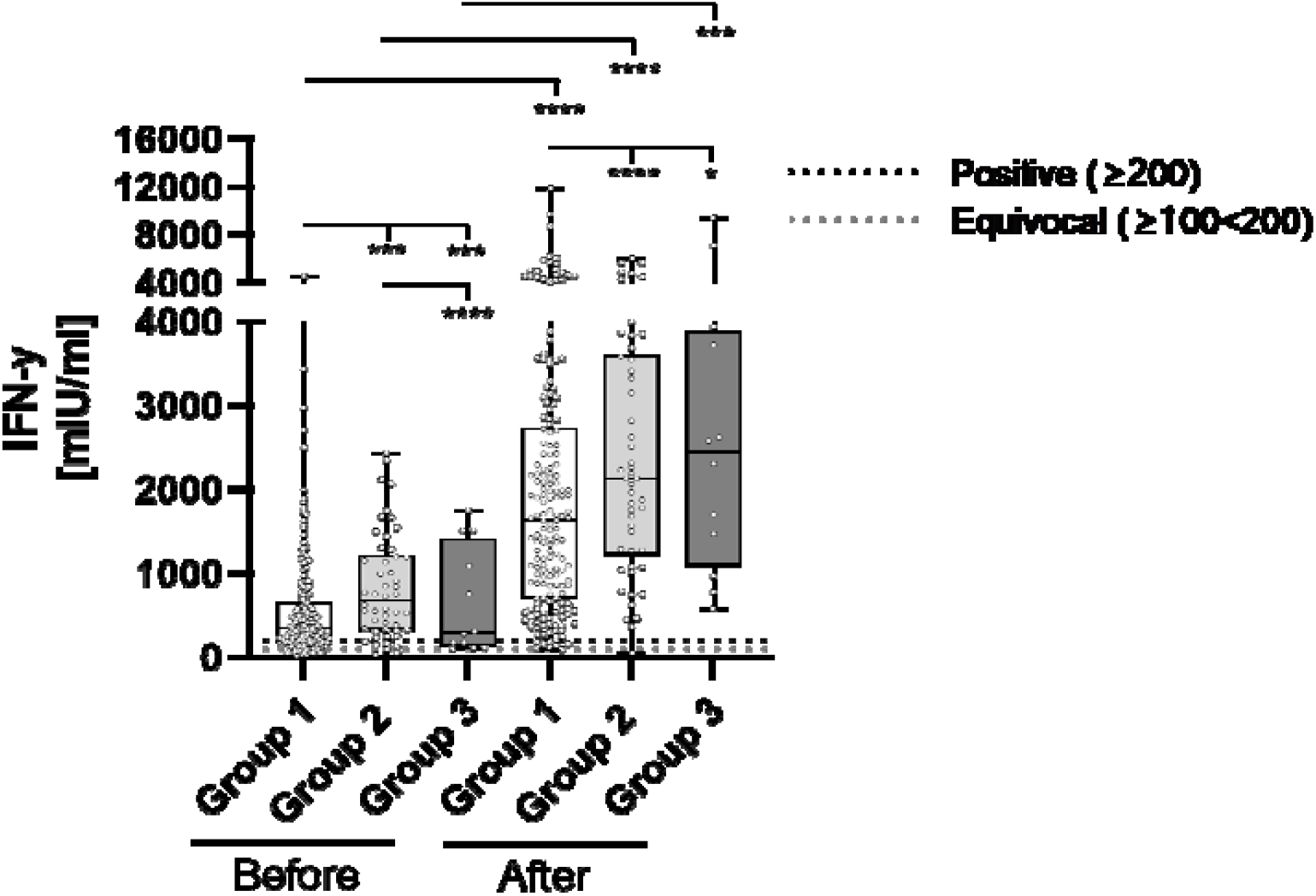
Comparative determination of SARS-CoV-2 specific t-cell response before and after third booster vaccination with BNT162b2. To evaluate differences between vaccination-strategies, participants were grouped into 3 cohorts: Group 1: three vaccine-doses of BNT162b2; Group 2: initially two vaccine-doses of ChAdOx1 and BNT162b2 booster-dosage; Group 3: heterologous vaccination-protocol (ChAdOx1+ BNT162b2) and BNT162b2 booster-dosage. Cellular immunity to SARS-CoV-2 was assessed by using an Interferon (IFN)-gamma release assay (IGRA) from Euroimmun (Quan-T-cell SARS-CoV-2 kit). Values > 200 mIU/ml were considered positive (horizontal black dotted line), whereas values between 100-200 mIU/ml were considered equivocal (horizontal gray dotted line). *p < 0.05; ***p < 0.001; ****p < 0.0001 (Mann-Whitney U-test).

#### Factors impacting the immune response

In the linear regression analysis, we identified the body mass index as a significant predictor for the antibody-level increase. The increase was also dependent on the antibody-level before booster. Neither sex, age or smoking had a significant effect. The estimated effect of a previous COVID-19 infection is negative, however since very few individuals (N=2) had an infection, no conclusion can be draws from this result. **Table 3** gives the result of the full regression model. The fit of the model was very good with an R^2^ value of 0.43. The regression coefficients remained virtually unchanged when only the significant variables, BMI and antibody-level before booster, remained in the final model (**Table 3**).

**Table 3:**
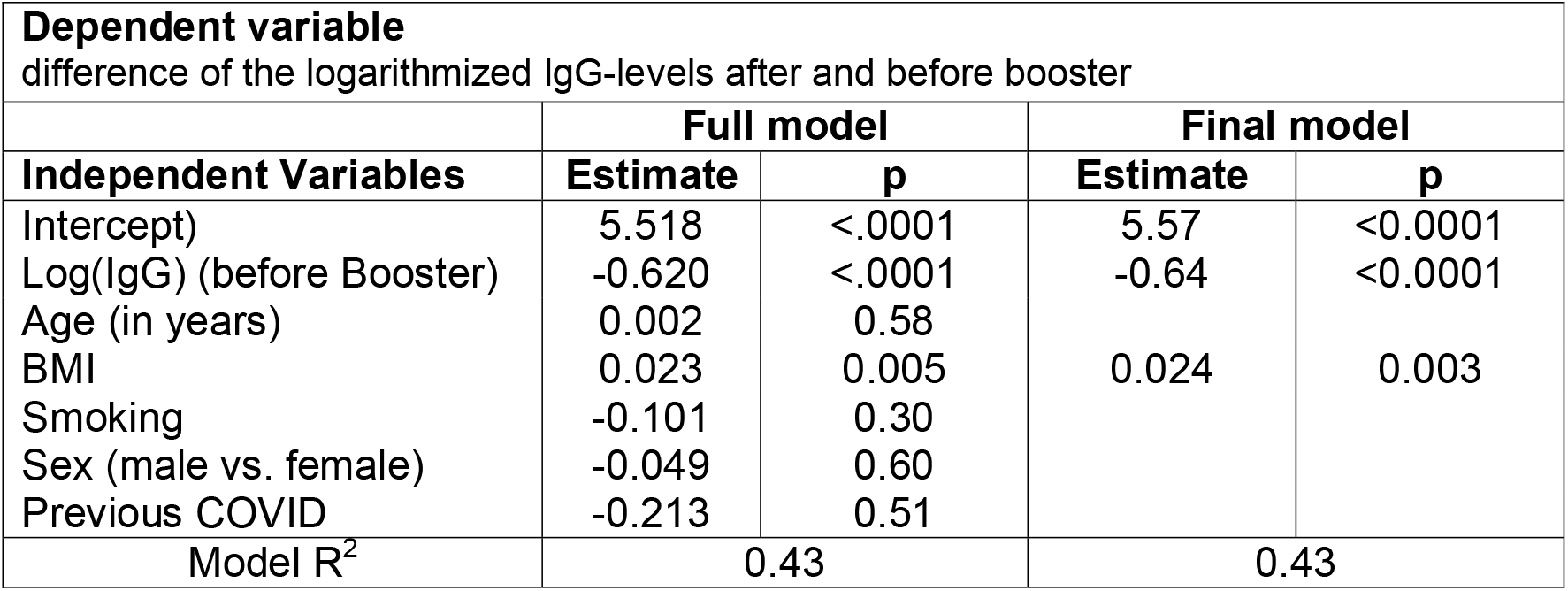
Results of the linear regression analysis

From the regression model, the estimated antibody-level showed both an absolute and relative increase given the BMI and the antibody-level before booster. **Table 4** provides the estimates for some selected values. For example, an individual with an antibody-level of 100 prior to booster and BMI of 25, has an estimated value 2552 after booster, which is an absolute increase of 2452 and 25.5-fold relative increase.

**Table 4:**
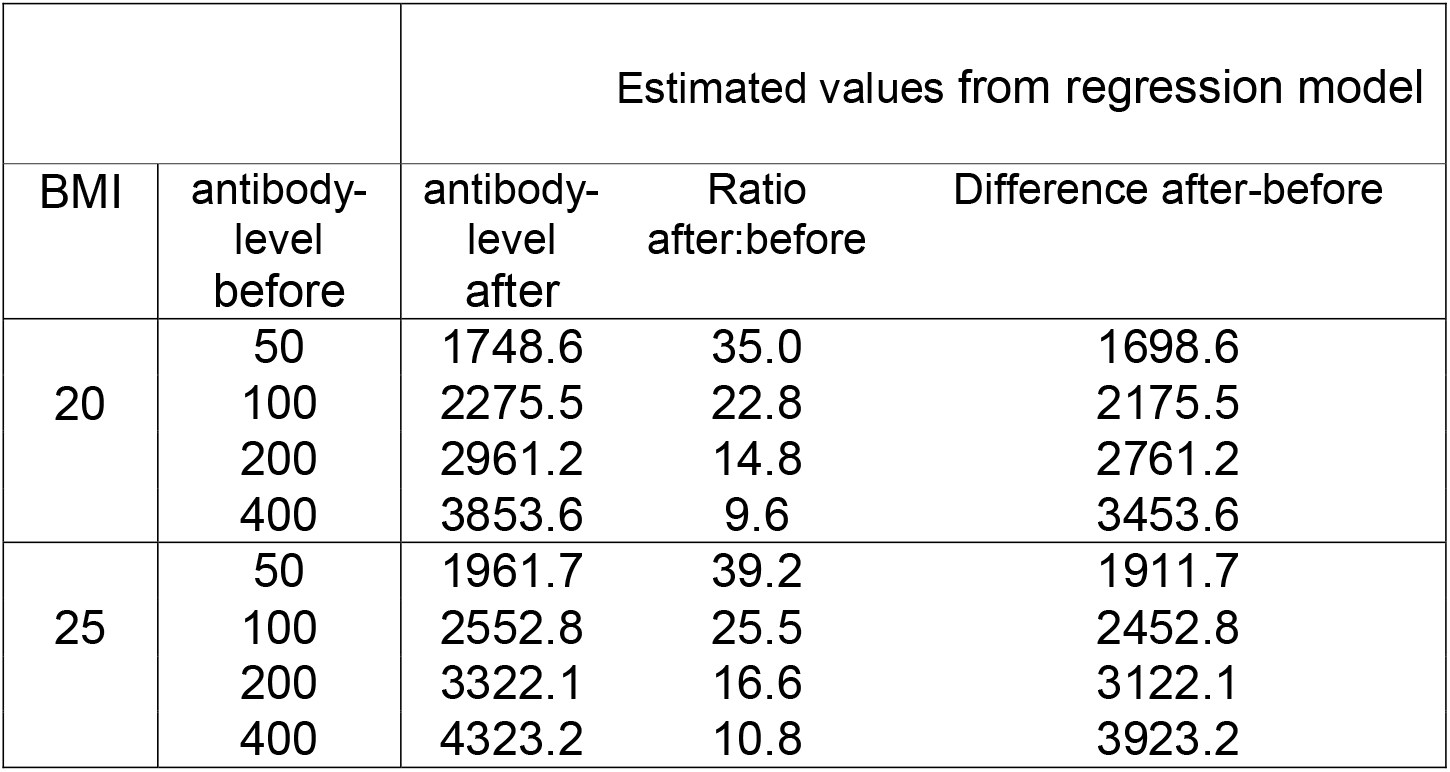
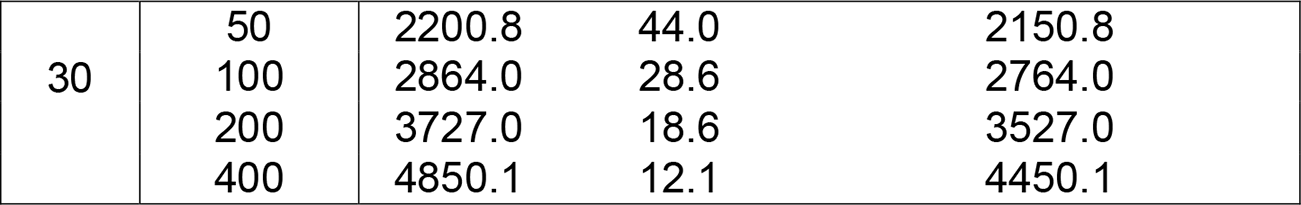
Estimates for different selected values.

|  |  | Estimated values from regression model |  |  |
| --- | --- | --- | --- | --- |
| BMI | antibody-level before | antibody-level after | Ratio after:before | Difference after-before |
| 20 | 50 | 1748.6 | 35.0 | 1698.6 |
|  | 100 | 2275.5 | 22.8 | 2175.5 |
|  | 200 | 2961.2 | 14.8 | 2761.2 |
|  | 400 | 3853.6 | 9.6 | 3453.6 |
| 25 | 50 | 1961.7 | 39.2 | 1911.7 |
|  | 100 | 2552.8 | 25.5 | 2452.8 |
|  | 200 | 3322.1 | 16.6 | 3122.1 |
|  | 400 | 4323.2 | 10.8 | 3923.2 |
| 30 | 50 | 2200.8 | 44.0 | 2150.8 |
|  | 100 | 2864.0 | 28.6 | 2764.0 |
|  | 200 | 3727.0 | 18.6 | 3527.0 |
|  | 400 | 4850.1 | 12.1 | 4450.1 |

#### Side effects of third dose

The majority of the study cohort reported some minor side-effect following the booster dose (**Table 1**). 54 participants (22.2%) reported no systemic or local reaction after administration of the third dose.

## Discussion

We present real-life data showing the impact of administration of a third dose of COVID-19 vaccination using BNT162b2 mRNA vaccine, following different vaccination protocols in a well-defined group of health care employees. Our data reveal a significantly increased cellular and humoral immune response to this additional dose.

Our data show that both humoral- and cellular immunity appear to be most persistent after double ChAdOx1 vaccination compared to the other vaccination strategies considered. In contrast, Dulovic et al. in their study showed that the persistence of neutralizing antibodies is shortest in individuals who have received the homologous ChAdOx1 vaccine. Contrarily, the administration of a heterologous- or a homologous mRNA strategy led to a stronger persistence. These discrepancies can be explained by the fact that Dulovic et al. only examined persistence over a maximum period of 65 days following administration of the second vaccine dose [19]. In our study, we considered a much longer time period of 6 months.

Our study shows the highest increase in INF-gamma release, binding-antibody expression, as well as neutralizing antibody capability for participants initially vaccinated two times with ChAdOx1. However, these participants showed the weakest humoral immune response before administration of the booster dose. A regression analysis detected a high BMI to be associated with an increased immune response.

Several studies have analyzed the effect of a third dose on the immune response and on the occurrence of infections, and showed an overall beneficial effect of the third dose, without relevant adverse events [6,20–22].

Atmar et al. analyzed in their phase 1-2 open-label clinical trial the efficiency of homologous and heterologous protocols for the third dose, and found a stronger increase in immunity against SARS-CoV-2 following a heterologous third dose [23]. This is also shown in the data presented here.

Besides the determined overall binding antibody titers, the neutralizing antibodies play a main role in the humoral immune response and are correlated with protection against severe COVID-19 outcome [24].

Using the commercial ELISA-based NeutraLISA assay from Euroimmun, our data show a substantial increase in neutralizing antibody capability in all participants after booster-administration. This is in line with the data presented from Atmar et al.[23]. Data from US and Israel underline the protective effect of a third dose of BNT162b2 against a severe COVID-19-course [25–27].

### Factors causing a reduced immune response

Multiple studies have tried, and are still attempting, to evaluate possible factors associated with a reduced immune response or with a complete non-response to anti-SARS-CoV-2-vaccination.

Within the study cohort presented here, smoking, increased age and BMI have previously been shown to have a negative impact in studies involving two doses of BNT162b2[28] or 9 months after vaccination, respectively [2]. This could not be seen in the results presented here (Table 3). In contrast, a higher BMI was associated with a stronger increase of the antibody titer after the third dose. This may be due to a lower antibody titer before administration of the third dose as reported previously [2]. Soffer et al. also found a positive correlation after COVID-19 infection for antibody titers and BMI, whereas Frasca et al. found the opposite correlation [29].

Other factors potentially influencing the immune response after a third dose of BNT162b2 are immunosuppression, for instance in patients after solid-organ transplantation [20].

In our cohort, we could not detect any negative impact of older age, although elderly participants are relatively underrepresented in this trial. Data published by Eliakim-Raz et. al. also described no correlation to higher age in a group older than 60 years [21].

Further studies are required to provide more accurate information on risk factors associated with reduced immunity to SARS-CoV-2.

#### Limitation

Despite the fact this study benefits from its real-life data in an important group of health-care workers, and a relatively large number of participants, it comes with some limitations.

This study is limited by its single-center design. The inclusion of health-care workers led to the overrepresentation of women and younger people, whereas groups with a higher risk for low immune response to vaccination, such as elderly and participants with an immunomodulatory treatment, are underrepresented. Unfortunately, the number of individuals in each group of vaccination protocols were unequally distributed, as there was no individual choice regarding the vaccine received.

## Conclusion

This study showed a BMI-dependent increase after the third dose of BNT162b2 following different vaccination protocols, although all participants showed a significant increase in their immune response. This effect, in combination with mild-to-none post-vaccination symptoms, underlines the potential beneficial effect of a BNT162b2-booster dose following 2 doses anti-SARS-CoV-2-vaccine. Further studies are needed to evaluate thresholds for the immune response – both humoral and cellular – and to detect the longevity of the booster-induced immunity.

## Data Availability

All data produced in the present study are available upon reasonable request to the authors.

## Acknowledgments

We would like to thank all team members at the Krankenhaus Reinbek St. Adolf-Stift, and all participating laboratories that analyzed these additional samples in a phase of high workload.

Thanks also to Rebecca Zimmer for her intense and rapid assistance with linguistic enrichment.

## References

1. Saciuk Y, Kertes J, Shamir Stein N, Ekka Zohar A. Effectiveness of a third dose of BNT162b2 mRNA vaccine. J Infect Dis [Internet]. 2021 Nov 2 [cited 2022 Jan 5]; Available from: http://www.ncbi.nlm.nih.gov/pubmed/34726239

2. Herzberg J, Fischer B, Lindenkamp C, Becher H, Becker A-K, Honarpisheh H, et al. Persistence of Immune Response in Health Care Workers After Two Doses BNT162b2 in a Longitudinal Observational Study. Front Immunol [Internet]. 2022 Mar 4 [cited 2022 Mar 10];13:701. Available from: https://www.frontiersin.org/articles/10.3389/fimmu.2022.839922/full

3. Levin EG, Lustig Y, Cohen C, Fluss R, Indenbaum V, Amit S, et al. Waning Immune Humoral Response to BNT162b2 Covid-19 Vaccine over 6 Months. N Engl J Med [Internet]. 2021 Dec 9 [cited 2022 Jan 5];385(24):e84. Available from: http://www.nejm.org/doi/10.1056/NEJMoa2114583

4. Hong E, Terrade A, Denizon M, Aouiti-Trabelsi M, Falguières M, Taha M-K, et al. Haemophilus influenzae type b (Hib) seroprevalence in France: impact of vaccination schedules. BMC Infect Dis [Internet]. 2021 Jul 30 [cited 2022 Jan 6];21(1):715. Available from: http://www.ncbi.nlm.nih.gov/pubmed/34330228

5. Bar-On YM, Goldberg Y, Mandel M, Bodenheimer O, Freedman L, Kalkstein N, et al. Protection of BNT162b2 Vaccine Booster against Covid-19 in Israel. N Engl J Med [Internet]. 2021 Oct 7 [cited 2022 Jan 3];385(15):1393–400. Available from: http://www.nejm.org/doi/10.1056/NEJMoa2114255

6. Barda N, Dagan N, Cohen C, Hernán MA, Lipsitch M, Kohane IS, et al. Effectiveness of a third dose of the BNT162b2 mRNA COVID-19 vaccine for preventing severe outcomes in Israel: an observational study. Lancet [Internet]. 2021 [cited 2022 Jan 3];398:2093–100. Available from: https://doi.org/10.1016/

7. Ducloux D, Colladant M, Chabannes M, Yannaraki M, Courivaud C. Humoral response after 3 doses of the BNT162b2 mRNA COVID-19 vaccine in patients on hemodialysis. Kidney Int [Internet]. 2021 Sep 1 [cited 2022 Jan 3];100(3):702–4. Available from: http://www.ncbi.nlm.nih.gov/pubmed/34216675

8. Food and Drug Administration. FDA Authorizes Booster Dose of Pfizer-BioNTech COVID-19 Vaccine for Certain Populations | FDA [Internet]. [cited 2022 Jan 5]. Available from: https://www.fda.gov/news-events/press-announcements/fda-authorizes-booster-dose-pfizer-biontech-covid-19-vaccine-certain-populations

9. European Medicines Agancy. Comirnaty and Spikevax: EMA recommendations on extra doses and boosters [Internet]. 2021. Available from: https://www.ema.europa.eu/en/news/comirnaty-spikevax-ema-recommendations-extra-doses-boosters

10. Ignoring WHO call, major nations stick to vaccine booster plans | Reuters [Internet]. [cited 2022 Jan 5]. Available from: https://www.reuters.com/world/europe/french-president-macron-third-covid-vaccine-doses-likely-elderly-vulnerable-2021-08-05/

11. Kamar N, Abravanel F, Marion O, Couat C, Izopet J, Del Bello A. Three Doses of an mRNA Covid-19 Vaccine in Solid-Organ Transplant Recipients. N Engl J Med [Internet]. 2021 [cited 2022 Jan 5];385(7):661–2. Available from: http://www.ncbi.nlm.nih.gov/pubmed/34161700

12. Munro APS, Janani L, Cornelius V, Aley PK, Babbage G, Baxter D, et al. Articles Safety and immunogenicity of seven COVID-19 vaccines as a third dose (booster) following two doses of ChAdOx1 nCov-19 or BNT162b2 in the UK (COV-BOOST): a blinded, multicentre, randomised, controlled, phase 2 trial. Lancet [Internet]. 2021 [cited 2021 Dec 4]; Available from: https://doi.org/10.1016/S0140-6736

13. Liu X, Shaw RH, Stuart AS V, Greenland M, Aley PK, Andrews NJ, et al. Safety and immunogenicity of heterologous versus homologous prime-boost schedules with an adenoviral vectored and mRNA COVID-19 vaccine (Com-COV): a single-blind, randomised, non-inferiority trial. Lancet (London, England) [Internet]. 2021 [cited 2022 Jan 6];398(10303):856–69. Available from: http://www.ncbi.nlm.nih.gov/pubmed/34370971

14. Shaw RH, Stuart A, Greenland M, Liu X, Van-Tam JSN, Snape MD, et al. Heterologous prime-boost COVID-19 vaccination: initial reactogenicity data. Lancet (London, England) [Internet]. 2021 [cited 2022 Jan 5];397(10289):2043. Available from: https://www.ncbi.nlm.nih.gov/pmc/articles/PMC8115940/

15. Tenbusch M, Schumacher S, Vogel E, Priller A, Held J, Steininger P, et al. Heterologous prime-boost vaccination with ChAdOx1 nCoV-19 and BNT162b2. Lancet Infect Dis [Internet]. 2021 [cited 2022 Jan 5];21(9):1212–3. Available from: http://www.ncbi.nlm.nih.gov/pubmed/34332707

16. Reindl-Schwaighofer R, Heinzel A, Mayrdorfer M, Jabbour R, Hofbauer TM, Merrelaar A, et al. Comparison of SARS-CoV-2 Antibody Response 4 Weeks After Homologous vs Heterologous Third Vaccine Dose in Kidney Transplant Recipients. JAMA Intern Med [Internet]. 2021 Dec 20 [cited 2022 Jan 5]; Available from: https://jamanetwork.com/journals/jamainternalmedicine/fullarticle/2787200

17. Herzberg J, Vollmer T, Fischer B, Becher H, Becker A-K, Sahly H, et al. A Prospective Sero-epidemiological Evaluation of SARS-CoV-2 among Health Care Workers in a German Secondary Care Hospital. Int J Infect Dis [Internet]. 2020 Oct 16 [cited 2020 Nov 18];0(0). Available from: http://www.ncbi.nlm.nih.gov/pubmed/33075538

18. Robert-Koch-Institut. Epidemiologisches Bulletin 43/2021. Epidemiol Bull. 2021;43.

19. Dulovic A, Kessel B, Harries M, Becker M, Ortmann J, Griesbaum J, et al. Comparative magnitude and persistence of SARS-CoV-2 vaccination responses on a population level in Germany. medRxiv [Internet]. 2021 Dec 5 [cited 2022 Feb 20];2021.12.01.21266960. Available from: https://www.medrxiv.org/content/10.1101/2021.12.01.21266960v1

20. Benotmane I, Gautier G, Perrin P, Olagne J, Cognard N, Fafi-Kremer S, et al. Antibody Response After a Third Dose of the mRNA-1273 SARS-CoV-2 Vaccine in Kidney Transplant Recipients With Minimal Serologic Response to 2 Doses. JAMA [Internet]. 2021 Sep 21 [cited 2022 Feb 4];326(11):1063. Available from: https://jamanetwork.com/journals/jama/fullarticle/2782538

21. Eliakim-Raz N, Leibovici-Weisman Y, Stemmer A, Ness A, Awwad M, Ghantous N, et al. Antibody Titers Before and After a Third Dose of the SARS-CoV-2 BNT162b2 Vaccine in Adults Aged ≥60 Years. JAMA [Internet]. 2021 Dec 7 [cited 2022 Feb 4];326(21):2203. Available from: https://jamanetwork.com/journals/jama/fullarticle/2786096

22. Demonbreun AR, Sancilio A, Vaught LA, Reiser NL, Pesce L, McNally EM, et al. Antibody titers before and after booster doses of SARS-CoV-2 mRNA vaccines in healthy adults. medRxiv [Internet]. 2021 Nov 21 [cited 2022 Feb 8];2021.11.19.21266555. Available from: https://www.medrxiv.org/content/10.1101/2021.11.19.21266555v1.full.pdf+html

23. Atmar RL, Lyke KE, Deming ME, Jackson LA, Branche AR, El Sahly HM, et al. Homologous and Heterologous Covid-19 Booster Vaccinations. N Engl J Med [Internet]. 2022 Jan 26 [cited 2022 Feb 8]; Available from: http://www.nejm.org/doi/10.1056/NEJMoa2116414

24. Cromer D, Steain M, Reynaldi A, Schlub TE, Wheatley AK, Juno JA, et al. Neutralising antibody titres as predictors of protection against SARS-CoV-2 variants and the impact of boosting: a meta-analysis. The Lancet Microbe [Internet]. 2022 Jan [cited 2022 Feb 8];3(1):e52–61. Available from: http://www.ncbi.nlm.nih.gov/pubmed/34806056

25. Barda N, Dagan N, Cohen C, Hernán MA, Lipsitch M, Kohane IS, et al. Effectiveness of a third dose of the BNT162b2 mRNA COVID-19 vaccine for preventing severe outcomes in Israel: an observational study. Lancet (London, England) [Internet]. 2021 Dec 4 [cited 2022 Feb 8];398(10316):2093–100. Available from: http://www.ncbi.nlm.nih.gov/pubmed/34756184

26. Patalon T, Gazit S, Pitzer VE, Prunas O, Warren JL, Weinberger DM. Odds of Testing Positive for SARS-CoV-2 Following Receipt of 3 vs 2 Doses of the BNT162b2 mRNA Vaccine. JAMA Intern Med [Internet]. 2022 Feb 1 [cited 2022 Feb 8];182(2):179. Available from: https://jamanetwork.com/journals/jamainternalmedicine/fullarticle/2786890

27. Perez JL, Pfizer MA. Breakthroughs that change patients’ lives Efficacy &amp; Safety of BNT162b2 booster-C4591031 2 month interim analysis [Internet]. [cited 2022 Feb 8]. Available from: https://www.cdc.gov/vaccines/acip/meetings/downloads/slides-2021-11-19/02-COVID-Perez-508.pdf

28. Herzberg J, Vollmer T, Fischer B, Becher H, Becker A-K, Honarpisheh H, et al. SARS-CoV-2-antibody response in health care workers after vaccination or natural infection in a longitudinal observational study. Vaccine [Internet]. 2021 Dec 3 [cited 2021 Dec 3]; Available from: https://www.sciencedirect.com/science/article/pii/S0264410X21015553?via%3Dihub

29. Frasca D, Reidy L, Cray C, Diaz A, Romero M, Kahl K, et al. Influence of obesity on serum levels of SARS-CoV-2-specific antibodies in COVID-19 patients. Ansari AA, editor. PLoS One [Internet]. 2021 Mar 24 [cited 2022 Feb 8];16(3):e0245424. Available from: https://dx.plos.org/10.1371/journal.pone.0245424

